# MRI connectivity-based spread of microglial activation in early Alzheimer’s disease

**DOI:** 10.1101/2022.02.22.22271354

**Authors:** Boris-Stephan Rauchmann, Matthias Brendel, Nicolai Franzmeier, Lena Trappmann, Mirlind Zaganjori, Estrella Morenas-Rodriguez, Selim Guersel, Lena Burow, Carolin Kurz, Jan Haeckert, Maia Tatò, Julia Utecht, Boris Papazov, Oliver Pogarell, Daniel Janowitz, Katharina Buerger, Michael Ewers, Carla Palleis, Endy Weidinger, Gloria Biechele, Sebastian Schuster, Anika Finze, Florian Eckenweber, Rainer Rupprecht, Axel Rominger, Oliver Goldhardt, Timo Grimmer, Daniel Keeser, Sophia Stoecklein, Olaf Dietrich, Peter Bartenstein, Johannes Levin, Günter Höglinger, Robert Perneczky

## Abstract

**Background:** Alzheimer’s disease (AD) is characterized by amyloid-β (Aβ) plaques, neurofibrillary tau tangles and neuroinflammation leading to brain functional connectivity changes and cognitive decline. There is evidence, that microglial activity is increased in AD and cognitive decline. Aβ and tau pathology appear to spread along pathways of highly connected brain regions, but it remains elusive if microglial activation follows a similar distribution pattern.

**Methods:** Thirty-two early AD subjects and 18 age-matched healthy cognitively normal controls were included from the prospective ActiGliA study. Differences between the diagnostic groups were explored for translocator protein (TSPO) positron emission tomography (PET) microglial activation, diffusion tensor imaging (DTI) structural connectivity and magnetic resonance imaging (MRI) functional connectivity. Associations between PET microglial activation with cognitive impairment, dementia severity and MRI connectivity measures were investigated within the diagnostic groups.

**Results:** AD patients showed increased TSPO PET tracer uptake bilaterally in the parahippocampal region compared to cognitively normal controls. Higher TSPO PET was associated with cognitive impairment and dementia severity in a disease stage dependent fashion. Inter-regional covariance in TSPO PET and standardized uptake value ratio (SUVR) was found to be preferentially distributed along functionally highly connected brain regions, with MRI structural connectivity showing a weaker association with microglial activation.

**Conclusion:** Neuroinflammation in AD is associated with clinical disease presentation, and like tau pathology, microglial activation seems to spread preferentially along highly connected brain regions. These findings support the important role of microglia in neurodegeneration and suggest that disease spreading throughout the brain along vulnerable connectivity pathways could guide future interventional anti-inflammatory therapy approaches to prevent disease progression.

## Introduction

Extracellular amyloid-β (Aβ) plaques and intracellular neurofibrillary tau tangles are the pathological hallmarks of Alzheimer’s disease (AD), leading to a cascade of brain changes resulting in cognitive decline and dementia (<u>Hardy and Higgins 1992</u>). Recent genetic, molecular and clinical evidence suggests that neuroimmune mechanisms are associated with AD risk and contribute to disease progression (Calsolaro and Edison 2016; Heneka et al. 2015). Patients with symptomatic AD show alterations in cerebrospinal fluid (CSF) and blood pro- and anti-inflammatory proteins (<u>Rauchmann et al. 2020; Rauchmann et al. 2019</u>), including the soluble triggering receptor expressed on myeloid cells 2 (sTREM2), a marker of activated microglia (Shen et al. 2019), which typically surround Aβ plaques as the brain’s main innate immune response (<u>Itagaki et al. 1989</u>). This suggests that neuroinflammation and microglial activation plays a key role in the pathogenesis of AD.

In vivo, microglial activation can be measured with positron emission tomography (PET) tracers targeting the 18 kDa translocator protein (TSPO), located on the outer mitochondrial membrane and overexpressed on activated immune cells (Edison and Brooks 2018). Increased microglial activation is apparent on TSPO PET scans in different neurodegenerative disorders, including AD, corticobasal syndrome and progressive supranuclear palsy (Heneka et al. 2015; Knezevic and Mizrahi 2018; Palleis et al. 2021).

Neuroinflammation, extracellular Aβ plaques and intracellular tau neurofibrils seem to be linked closely. Within the microglia population, two distinct expression profiles were recently identified, associated with either Aβ load or tissue phosphorylated tau (ptau) (<u>Gerrits et al. 2021</u>). Colocalization of tau and activated microglia was demonstrated (Hayes et al. 2002; Dani et al. 2018), and Aβ plaque-dependent microglia was identified (Plescher et al. 2018). Furthermore, TSPO PET binding was shown to correlate with increasing Aβ accumulation in AD mouse models (Blume et al. 2018), and microglial activation may also act as a link between Aβ and tau pathology (Kitazawa et al. 2004).

Several studies in AD have demonstrated that changes in functional magnetic resonance imaging (fMRI) connectivity are associated with hallmark protein aggregations. Highly connected hub regions show increased tau accumulation on PET imaging, accelerated by Aβ pathology (Franzmeier et al. 2020; Vogel et al. 2020), supporting a model of transneuronal spread of tau molecules (Cope et al. 2018). Increasing evidence indicates that the spread of tau tangles in the neocortex is accompanied by microglial activation (Sheffield et al. 2000; Hopp et al. 2018; Dani et al. 2018), with both types of pathology seemingly following a Braak-stage-like distribution pattern along an activated network of microglia, suggesting that Aβ and microglia together initiate tau spreading across the brain (Pascoal et al. 2021; Braak et al. 2006).

The main aims of the present study were to characterize the pattern of TSPO-PET microglial activation in early AD stages vs cognitively normal controls, to explore the associations between activated microglia and clinical endpoints, and to identify the brain structural and functional network connectivity correlates of the activated immune system. We used data from the prospective Activity of Cerebral Networks, Amyloid and Microglia in Aging and Alzheimer’s disease (ActiGliA) study to address our hypotheses.

## Methods

### Study design and participants

The data used in this study originate from the baseline dataset of the ActiGliA study, a prospective, longitudinal, observational, single-center study of the Munich Cluster for Systems Neurology (SyNergy) at Ludwig-Maximilians-University (LMU) Munich, initiated in 2017. Participants were recruited through specialized outpatient clinics at the LMU hospital Department of Psychiatry and Psychotherapy, Department of Neurology and Institute of Stroke and Dementia Research and the Department of Psychiatry and Psychotherapy of the Technical University Munich. ActiGliA comprises comprehensive neurocognitive, clinical and lifestyle assessments based on the German Center for Neurodegenerative Disorders (DZNE)-Longitudinal Cognitive Impairment and Dementia (DELCODE) study (Jessen et al.

2018); MRI and PET imaging using tracers for Aβ and TSPO; and fluid biobanking, including CSF, plasma, serum, saliva, DNA, RNA and peripheral blood mononuclear cells. Patients with early AD (subjective cognitive impairment, MCI and mild AD dementia) and corticobasal syndrome (CBS) (Schmitt et al. 2021) and age-matched cognitively normal controls were included after providing written informed consent in line with the declaration of Helsinki. The study was approved by the ethics committee of LMU Munich (project numbers 17-755 and 17-569).

Out of 140 ActiGliA participants, all consecutive cases meeting the inclusion criteria for the present analyses were included, resulting in a cohort of N=32 early AD patients and N=18 cognitively normal controls. N=90 participants were not considered for the present analysis due to a diagnosis of N=50 CBS or other non-AD disorder (N=23). N=16 participants were excluded because of missing neuropsychological test results and N=1 participant was excluded because of extensive white matter lesions (**Figure 1**). Cognitive normal controls (CN) were defined as participants without cognitive impairment (Clinical Dementia Rating (CDR) global score = 0, Consortium to Establish a Registry for AD neuropsychological battery (CERAD-NB) total score ≥ 69) (Chandler et al. 2005) and no indication of Aβ pathology on PET (negative visual read) and/or CSF examination (normal Aβ42/40-ratio as defined below). AD continuum was defined as CDR global score ≥ 0.5, CERAD-NB total score ≤ 84 and presence of Aβ pathology on PET and/or CSF examination.

**Figure 1:**
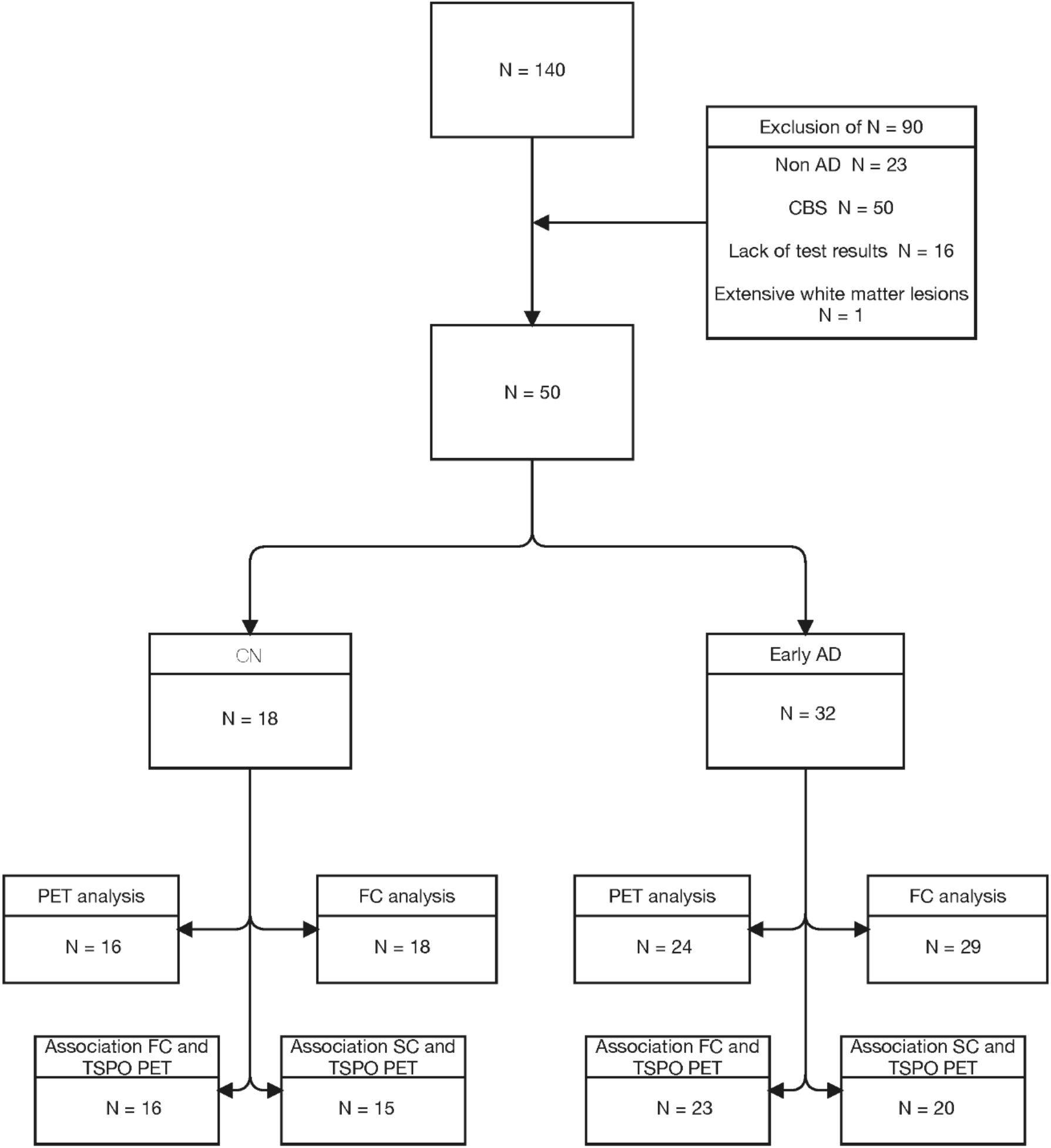
Flow chart showing the number and reasons for study exclusions. Abbreviations: AD, Alzheimer’s disease; CN, cognitively normal; CBS, Corticobasal syndrome; FC, functional connectivity; SC, structural connectivity; TSPO, mitochondrial translocator protein.

### Clinical assessments

The CDR, CERAD-NB (Fillenbaum et al. 2008) and Mini-Mental State Examination (MMSE) (<u>Folstein et al. 1975</u>) were conducted by trained psychologists at the LMU hospital memory clinic. Using the CERAD-NB battery a total score was created as shown previously, comprising the six sub-tests semantic fluency (animals/60 seconds), modified Boston Naming Test, Word List Learning, Constructional Praxis, Word List Recall and Word List Recognition Discriminability, with higher scores indicating better performance (Chandler et al. 2005).

### Genetic polymorphisms

*TSPO* and apolipoprotein E (*APOE*) genotyping was performed at the Departments of Psychiatry and Psychotherapy of University of Regensburg and LMU Munich respectively. Genomic DNA was extracted from whole blood using a SQ Blood DNA kit from Omega Bio-Tek (Norcross, GA, USA) according to the manufacturer’s protocol. DNA quality was assessed by optical absorbance and gel electrophoresis. TaqMan quantitative polymerase chain reaction assays were used for amplification and Sanger method for sequencing.

Binding affinity of the [^18^F]GE-180 TSPO ligand is affected by the co-dominant rs6971 (Ala/Thr) single nucleotide polymorphisms (SNP) of the *TSPO* gene and needs to be considered in the imaging analysis (Kreisl et al. 2013). High-affinity binders (HAB) are Ala/Ala carriers, low-affinity binders (LAB) are Thr/Thr carriers and mixed-affinity binders (MAB) are Ala/Thr carriers. Only HAB and MAB carriers were included in the PET analyses, with N=6 LAB excluded. All TSPO PET analyses were adjusted for binding status. The two SNPs rs429358 and rs7412 defining the *APOE* ε2, ε3 and ε4 alleles were analyzed to determine the *APOE* genotype. Sequencing data were analyzed using SnapGene software (GSL Biotech; http://snapgene.com). Participants were dichotomized into carriers vs non-carriers of the ε4 allele.

### CSF analyses

CSF peptide measures were generated from aliquoted samples using commercially available (Fujirebio, Malvern, PA) enzyme-linked immunosorbent assays (ELISAs). Aβ positivity was defined as a CSF Aβ42/40-ratio of <5.5%, as suggested previously (Dumurgier et al. 2015). Concentrations of total tau (ttau) and ptau181 were measured in CSF using the Innotest htau-Ag, and Innotest P-tau (181P) ELISA assays (Fujirebio, Europe). The CSF sTREM2 concentrations were measured at the DZNE Munich based on previously established in-house ELISA protocols using duplicate samples (Suárez-Calvet et al. 2016). Average measurements of the duplicates were used for the statistical analyses after applying method and plate-specific correction procedures.

### MRI acquisition and preprocessing

MRI data for the entire ActiGliA cohort was acquired at the Department of Radiology of LMU Munich on a Siemens 3T Magnetom Skyra MR system (Siemens Healthineers, Erlangen, Germany). A 0.8 cm isovoxel high resolution T1-weighted structural MRI sequence (repetition time (TR), 2060 ms; echo time (TE), 2.17 ms; flip angle (FA), 12 deg; field of view (FoV), 240 mm), a diffusion weighted imaging (DWI) MRI sequence with a multi-band acceleration factor 3 (TR, 3800 ms; TE, 104.8 ms; b-value, 2000 s/mm^²^; 108 diffusion directions; FA, 90 deg, FoV, 240 mm) and a 2.5 mm, 10:32 min resting state fMRI sequence with multi-band acceleration factor 6 (TR, 780 ms; TE, 33 ms; FA, 50 deg; FoV, 210 mm) were acquired.

Resting state functional connectivity was analyzed using the CONN fMRI functional connectivity toolbox (ver. 17, www.nitrc.org/projects/conn) in MATLAB (MathWorks, Inc., Natick, Massachusetts, USA). Preprocessing comprised visual inspection, volume-based fMRI analyses, including realignment, slice-time correction, segmentation and structural and functional normalization, ART-based outlier detection and functional smoothing using a 6 mm kernel. The pipeline details can be found elsewhere (https://web.conn-toolbox.org/fmri-methods/preprocessing-pipeline).

All diffusion tensor imaging (DTI) images were visually inspected. Preprocessing was performed using ExploreDTI (Leemans et al. 2009), a MATLAB (MathWorks, Inc., Natick, Massachusetts, USA) toolbox, including motion correction and eddy current correction and correction for echo planar imaging (EPI) distortions.

### Positron emission tomography acquisition and preprocessing

Patients were scanned at the Department of Nuclear Medicine of LMU Munich on a Biograph 64 PET/CT scanner (Siemens Healthineers, Erlangen, Germany). Prior to PET acquisition a low-dose CT scan was acquired for attenuation correction. Emission data were acquired dynamically over 90 min or static from 60-80 min p.i., starting with the injection of 189 ± 12 MBq [^18^F]GE-180 as an intravenous bolus. The specific activity was on average 1.714 ± 523 GBq/μmol at the end of radiosynthesis, and the injected mass was 0.13 ± 0.05 nmol (based on N = 5 syntheses). Images were reconstructed using a 3-dimensional ordered subsets expectation maximization algorithm (16 iterations, 4 subsets, 4 mm gaussian filter) with a matrix size of 336 × 336 × 109, and a voxel size of 1.018 × 1.018 × 2.027 mm. Standard corrections for attenuation, scatter, decay and random counts were applied.

TSPO PET scans were co-registered to the subject’s individual T1-weighted structural MRI scan, skull-stripped using FSL BET. Non-linear spatial normalization parameters to the Montreal Neurological Institute (MNI) standard space were estimated using Advanced Normalization Tools (ANTs). The obtained spatial normalization parameter estimations were subsequently used to normalize the TSPO PET scan to the MNI space. To obtain standardized uptake value ratios (SUVRs), the TSPO PET scans were intensity normalized using the inferior cerebellar grey matter as reference region (Baker et al. 2017).

### Analysis of TSPO PET differences between diagnostic groups

Differences between the diagnostic groups were estimated in a voxel-wise analysis using SPM12 (The Wellcome Centre for Human Neuroimaging, London, UK). MNI normalized TSPO PET images were spatially smoothed using an 8 mm FWHM kernel. In an atlas-based analysis the individual TSPO tracer uptake was calculated in the regions of the Brainnetome atlas, a multimodal connectivity based parcellation framework atlas based on a 246-region brain parcellation, including 210 cortical and 36 subcortical subregions (Fan et al. 2016).

### Analysis of the structural and functional connectomes and their associations with TSPO PET covariance

The Brainnetome atlas was used to determine the individual in vivo connectivity architecture when assessing structural and functional connectivity. Group differences in functional and structural connectivity between AD and control subjects were assessed using the CONN toolbox and GraphVar (Kruschwitz et al. 2015). The associations between TSPO PET tracer uptake and functional and structural connectivity were analyzed using in-house MATLAB scripts for z-scaling, covariance calculation and linear regression analysis.

### Statistical analyses

Group differences in functional connectivity were tested in a general linear model using individual adjacency matrices of the Brainnetome atlas based 246 regions parcellation in a region of interest (ROI)-to-ROI analysis in the CONN toolbox. All results were adjusted for age, sex and education with false discovery rate (FDR) correction for multiple comparisons with a significance level of p<0.05. Differences in structural connectivity between the diagnostic groups were tested in a general linear model using the individual DTI derived adjacency matrices of the number of tracts comprising the 246 BN atlas-based regions in the GraphVar toolbox adjusted for age, sex and education. Associations between clinical scores and TSPO PET uptake was tested using a quadratic regression analysis in SPSS (IBM SPSS Statistics for Windows, Version 26.0. Armonk, NY: IBM Corp) assuming a disease stage-dependent microglial activation pattern, as shown previously (Blume et al. 2018).

To assess the association between MRI functional/structural connectivity and microglial activation on PET, the Fisher r-to-z transformed matrices of functional connectivity and the z-transformed DTI-derived number of tracts matrices for each subject were used. First, the Brainnetome atlas-based TSPO PET SUVR for each subject was concatenated, vectorized and z-transformed to remove inter-subject differences of the tracer uptake (Veronese et al. 2019), followed by a pairwise interregional correlation to calculate a TSPO PET covariance adjacency matrix for each diagnostic group. We vectorized the TSPO PET covariance matrix and the mean functional and structural connectivity adjacency matrices and performed a linear regression analysis to predict TSPO PET covariance from connectivity. Due to skewed distribution the vectorized structural connectivity was log-transformed for further analysis, not connected brain regions were excluded. Second, we tested how depending on the TSPO PET uptake in a seed region connectivity was associated with TSPO PET uptake in the target regions. To this end, we first calculated the mean adjacency matrix of functional and structural connectivity, followed by correlating each mean matrix with the TSPO SUVR vector to obtain a correlation vector. Subsequently, in a linear regression analysis we tested how the TSPO PET SUVRs were associated with the correlation vectors between functional/structural connectivity and TSPO SUVR. Finally, the association between TSPO PET uptake in functional/structural connectivity target regions and the functional/structural connectivity in hotspot and cold spot seed areas of TSPO PET uptake were assessed. The mean SUVR TSPO uptake was sorted discerningly in all 246 BN regions for each diagnostic group to define a hot and a cold spot region. Subsequently, in consequent regression analyses the TSPO PET uptake in all target regions was predicted by the functional/structural connectivity of the hotspot and cold spot.

## Results

Differences in sociodemographic and clinical data between the diagnostic groups are presented in **Table 1**. There were no differences in age, sex, weight and years of education. Compared to cognitively normal controls, AD subjects had significantly (p<0.001) worse cognitive performance (CERAD-NB and MMSE) and higher CDR-sob ratings. As expected, AD patients showed more pathological CSF biomarker concentrations compared to the control group (lower Aβ42 and Aβ42/40-ratio, higher ptau181 and ttau).

**Table 1:** Sociodemographic, clinical and biomarker characteristics of the cohort

| Characteristic | CN | AD patients | P-value |
| --- | --- | --- | --- |
| Age [years] (SD) | 69.22 (7.53) | 71.31 (7.52) | 0.35 |
| Sex | 10/18 (56%) woman | 18/32 (56%) woman | 0.60 |
| Weight [kg] (SD) | 76.39 (17.23) | 69.50 (13.78) | 0.13 |
| Education [years] (SD) | 14.83 (3.97) | 14.81 (4.13) | 0.99 |
| CDR sob (SD) | 0.06 (0.16) | 3.16 (1.73) | <0.001 |
| CERAD-NB (SD) | 86.42 (5.93) | 58.53 (14.05) | <0.001 |
| MMSE (SD) | 29.28 (1.07) | 24.25 (3.17) | <0.001 |
| CSF Aβ42 [pg/ml] (SD) | 1060.41 (482.93) | 465.96 (167.51) | <0.001 |
| CSF Aβ42/40-ratio (SD) | 7.48 (1.01) | 3.58 (0.96) | <0.001 |
| CSF ptau181 [pg/ml] (SD) | 49.75 (17.39) | 102.17 (46.78) | <0.001 |
| CSF ttau [pg/ml] (SD) | 230.37 (105.57) | 635.21 (606.34) | 0.009 |
| CSF sTREM2 [ng/ml] (SD) | 10.35 (3.00) | 11.31 (3.49) | 0.37 |
| TSPO PET SUVR (SD) | 0.94 (0.04) | 1.08 (0.08) | <0.001 |
| APOE ε4 allele carriers (SD) | 4/18 (22%) | 23/29 (79%) | <0.001 |
| rs6971 binding status (SD) | 15/18 (83%) MAB<br>3/18 (17%) HAB | 6/30 (20%) LAB<br>15/30 (50%) MAB<br>9/30 (30%) HAB | 0.041 |

Abbreviations: AD, Alzheimer's disease; CN, cognitively normal controls; CDR SoB,
Clinical Dementia Rating Sum of Boxes; CERAD-NB, Consortium to Establish a Registry for
Alzheimer's Disease neuropsychological battery total score; MMSE, Mini-Mental Status
Examination; Aβ42, amyloid-β42; Aβ -Ratio, Ratio Aβ42 to Aβ 40; ptau, phosphorylated tau
protein; ttau, total tau protein; sTREM2, soluble triggering receptor expressed on myeloid
cells 2; TSPO SUVR, mitochondrial translocator protein standardized uptake value; *APOE*,
Apolipoprotein E; LAB, low-affinity binder; MAB, mixed-affinity binder; HAB, high-affinity
binder.

### Group differences on PET and MRI

Supporting the view of increased microglial activation in AD, the AD patient group showed increased TSPO PET tracer uptake compared to cognitively normal controls (p<0.001) in the bilateral parahippocampal gyrus (**Table 2, Figure 2A**). Group differences in functional connectivity were detected on the whole brain level and within the default mode network comparing cognitively normal controls and AD patients. On the whole brain level, a widespread hypoconnectivity in AD with some hyperconnectivity mainly between temporal and cingulate/occipital and frontal brain regions was revealed (**Figure 2B)**. Within the default mode network hypoconnectivity between bilateral hippocampal regions was observed in AD (**Supplementary Figure 1)**. No significant differences were detected in the visual dorsal attention ventral attention limbic and frontotemporal networks. A widespread hyperconnectivity pattern was revealed in the structural connectivity analysis with partial hyperconnectivity mainly between temporooccipital and temporoparietal regions (**Figure 2C**).

**Table 2:** Regions with highest TSPO PET tracer uptake in Alzheimer’s disease compared with cognitively normal controls

|  | **Left hemisphere** | **Right hemisphere** |
| --- | --- | --- |
| **Atlas regions (Talairach)** | Parahippocampal Uncus<br>(Amygdala) | Parahippocampal Uncus<br>(Amygdala) |
| **Centroid (MNI Coordinates)** | x = -22, y = -8, z = -20 | x = 22, y = -2, z = -20 |
| **Voxels** | 34 | 20 |
| **Threshold (P-Value)** | 0.001 | 0.001 |
| **Voxel Threshold** | 20 | 20 |

**Figure 2:**
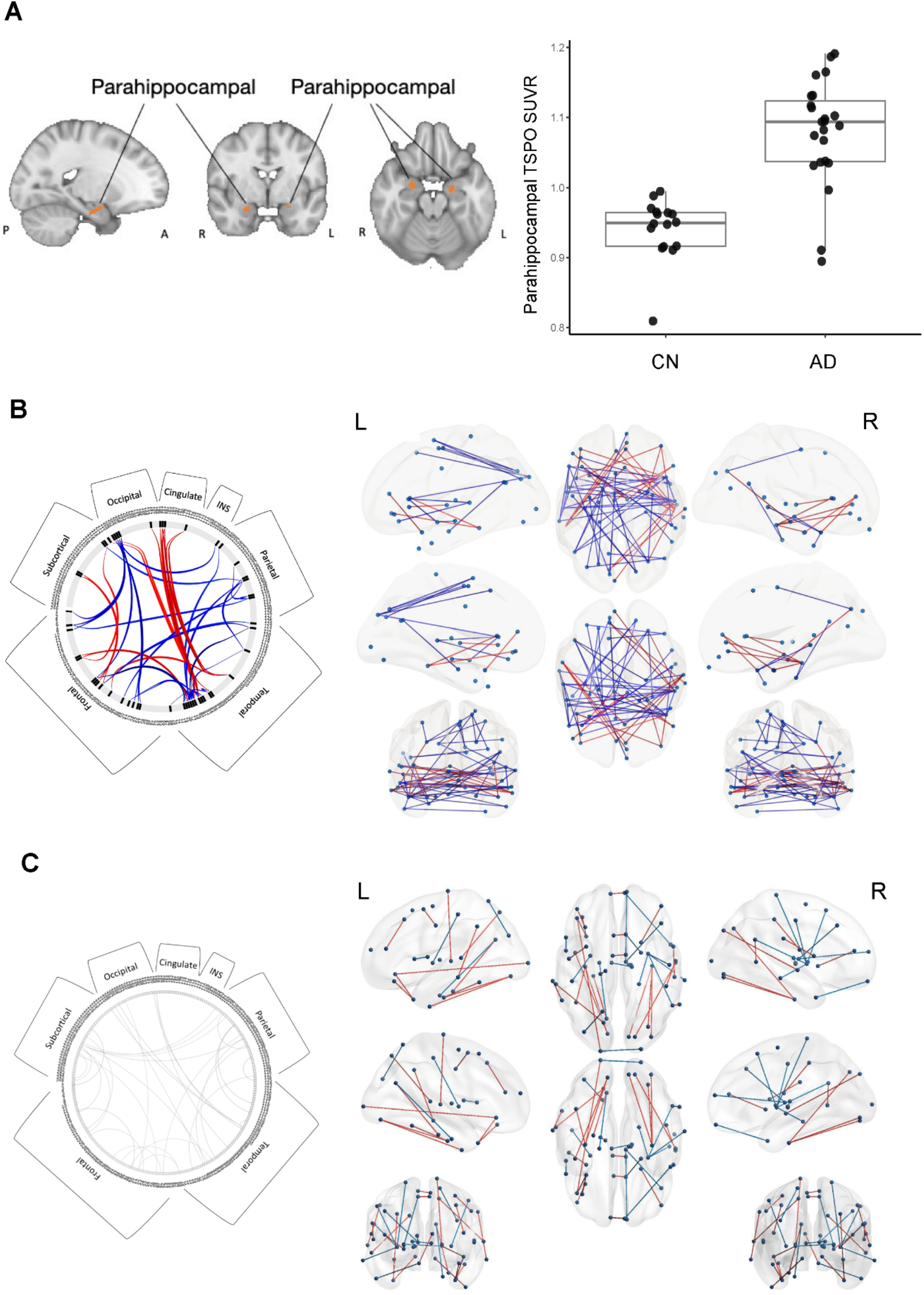
**(A)** Voxel wise group comparisons between Alzheimer’s disease and cognitively normal controls (p<0.001); scatterplot showing individual and mean differences between patients and cognitively normal controls in mitochondrial translocator protein standardized uptake value (SUVR). **(B)** Group differences in region of interest-based functional connectivity between Alzheimer’s disease and cognitively normal controls on whole brain level (cognitively normal controls <patients, FDR corrected p<0.05). **(C)** Differences in structural connectivity using diffusion tensor imaging-derived numbers of tracts on whole brain level (cognitively normal controls <patients, FDR corrected p<0.05). The numbers in the connectome ring in B) and C) correspond to regions in the Brainnetome atlas (**Supplementary Table 1)**. Abbreviations: AD, Alzheimer’s disease; CN, cognitively normal controls; TSPO, mitochondrial translocator protein; SUVR, standardized uptake value; L left; R, right.

### Associations between TSPO PET, dementia severity and cognitive performance

We analyzed how TSPO tracer uptake in the parahippocampal region was associated with dementia severity and cognitive impairment. Our analysis revealed a quadratic association, between parahippocampal TSPO PET SUVR and CDR sob (R^2^=0.39, p<0.001), CERAD-NB total score (R^2^=0.20, p<0.017) and MMSE score (R^2^=0.15, p=0.049) (**Figure 3)**.

**Figure 3:**
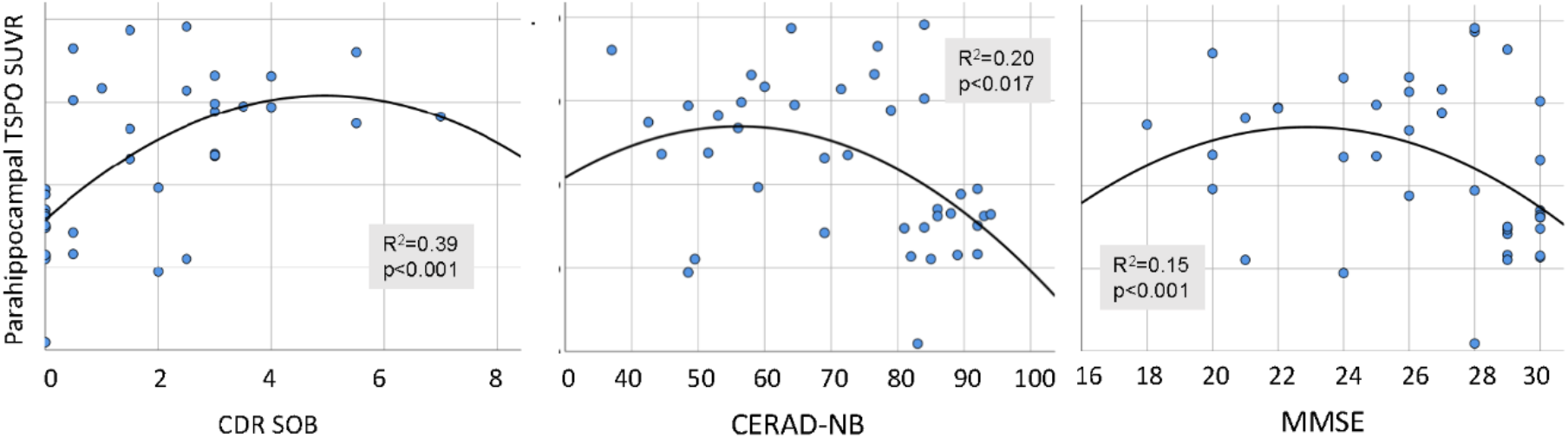
Associations between dementia severity and cognitive impairment and parahippocampal TSPO PET uptake in the whole study cohort, adjusted for age, sex, TSPO binding status and diagnostic group. Abbreviations: TSPO SUVR, mitochondrial translocator protein standardized uptake value; CDR SOB, Clinical Dementia Rating Sum of Boxes; CERAD-NB, Consortium to Establish a Registry for Alzheimer’s Disease neuropsychological battery total score; MMSE, Mini-Mental Status Examination.

### Associations between functional and structural connectivity and TSPO PET

We assessed how TSPO PET microglial activation is associated with functional and structural connectivity in AD patients and cognitively normal controls. Adjacency matrices of functional and structural connectivity using the 246 Brainnetome regions were generated for the control and AD patient groups (**Figure 4B** and **Figure 4C**). Using the same regions, TSPO PET derived covariance matrices were calculated for cognitively normal controls and AD patients (**Figure 4A)**. In a subsequent regression analysis, the associations between functional and structural connectivity with TSPO PET tracer uptake covariance were assessed. A significant association between functional connectivity and TSPO PET covariance was revealed in the AD (β=0.35, p<0.001) and in the control group (β=0.47, p<0.001) (**Figure 5A)**. For structural connectivity, a weak association with TSPO PET covariance was shown for cognitively normal controls (β =0.08; p<0.001) and for AD patients (β=0.06; p<0.001) (**Figure 5B)**. Log-transformed data were reported because of skewed distribution.

**Figure 4:**
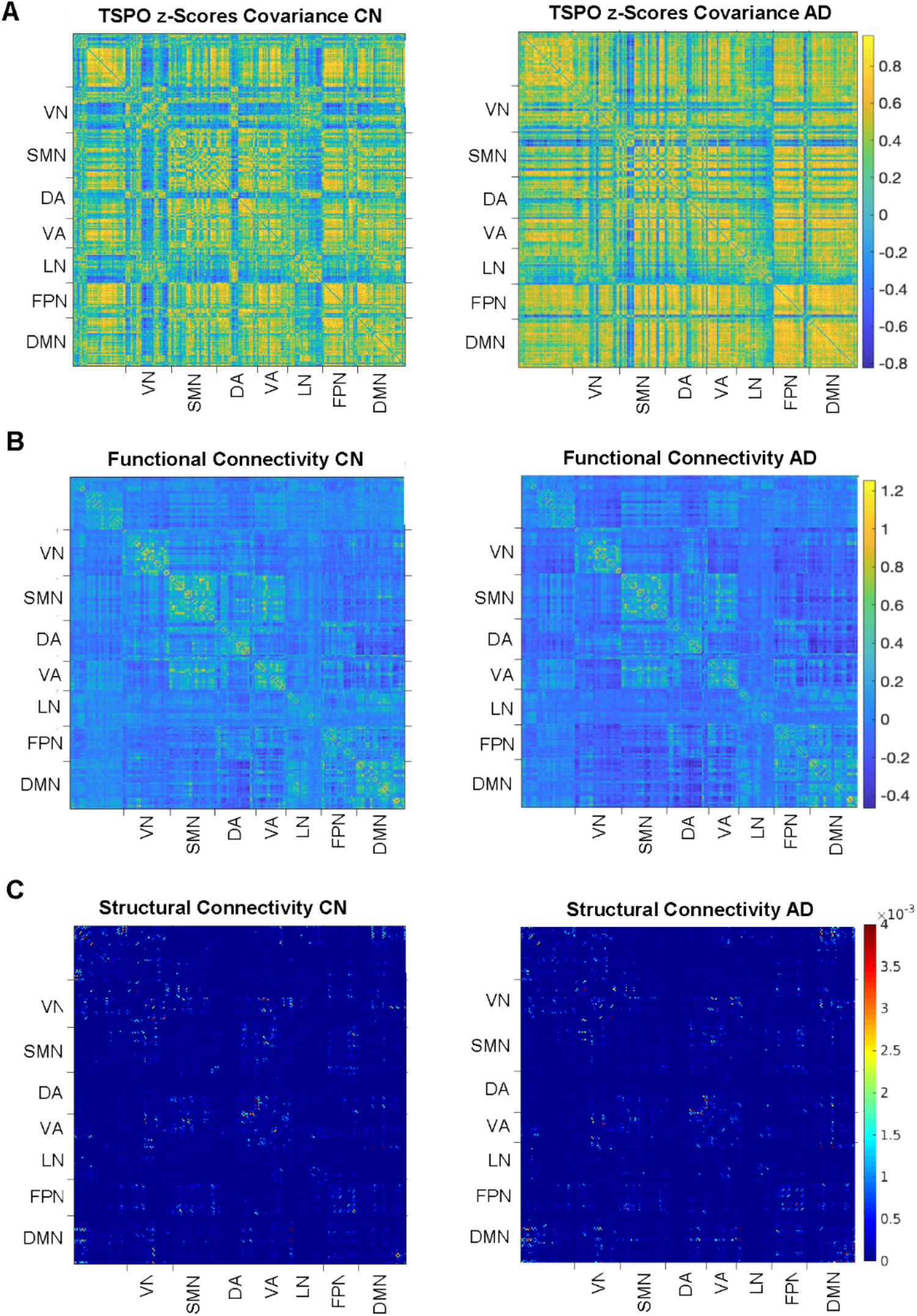
**(A)** Mean TSPO PET covariance matrix of cognitively normal controls and Alzheimer’s disease patients. **(B)** Mean adjacency matrix of functional connectivity **(C)** Mean adjacency matrix of structural connectivity. Abbreviations: AD, Alzheimer’s disease; CN, cognitively normal controls; TSPO, mitochondrial translocator protein; SUVR, standardized uptake value; L left; R, right; VN, visual network, SMN, sensorimotor network, DA, dorsal attention network; VA, ventral attention network; LN, limbic network; FPN, fronto parietal network; DMN, default mode network.

**Figure 5:**
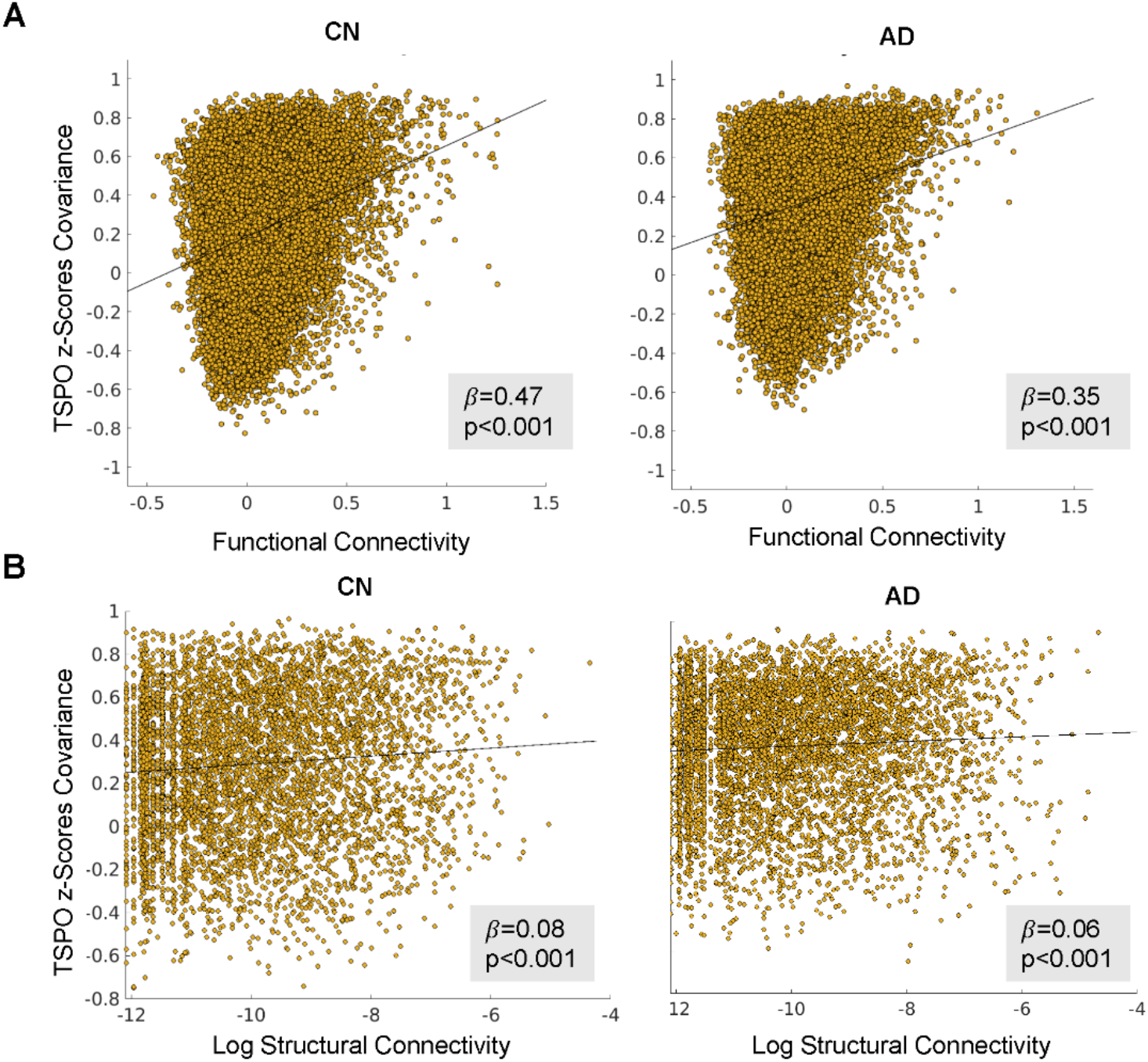
**(A)** Associations between functional connectivity and TSPO PET microglial activation. **(B)** Associations between structural connectivity and TSPO PET microglial activation. Abbreviations: AD, Alzheimer’s disease; CN, cognitively normal controls; TSPO, mitochondrial translocator protein; Log, logarithm.

### Network connectivity prediction of TSPO PET tracer uptake

We explored if TSPO PET tracer uptake in a particular region is predictive of the TSPO levels in a connected region, following a similar approach proposed recently for the association between functional connectivity and tau PET (Franzmeier et al. 2019). For seed regions with higher levels of TSPO PET uptake functional connectivity was associated with higher levels of TSPO PET uptake in the target regions and vice versa in the cognitively normal controls (β=0.57, p<0.001) and the AD group (β=0.83, p<0.001) (**Figure 6A)**. A somewhat weaker association was shown for structural connectivity in cognitively normal controls (β=0.37, p<0.001) and AD patients (β=0.36, p<0.001) (**Figure 6B)**.

**Figure 6:**
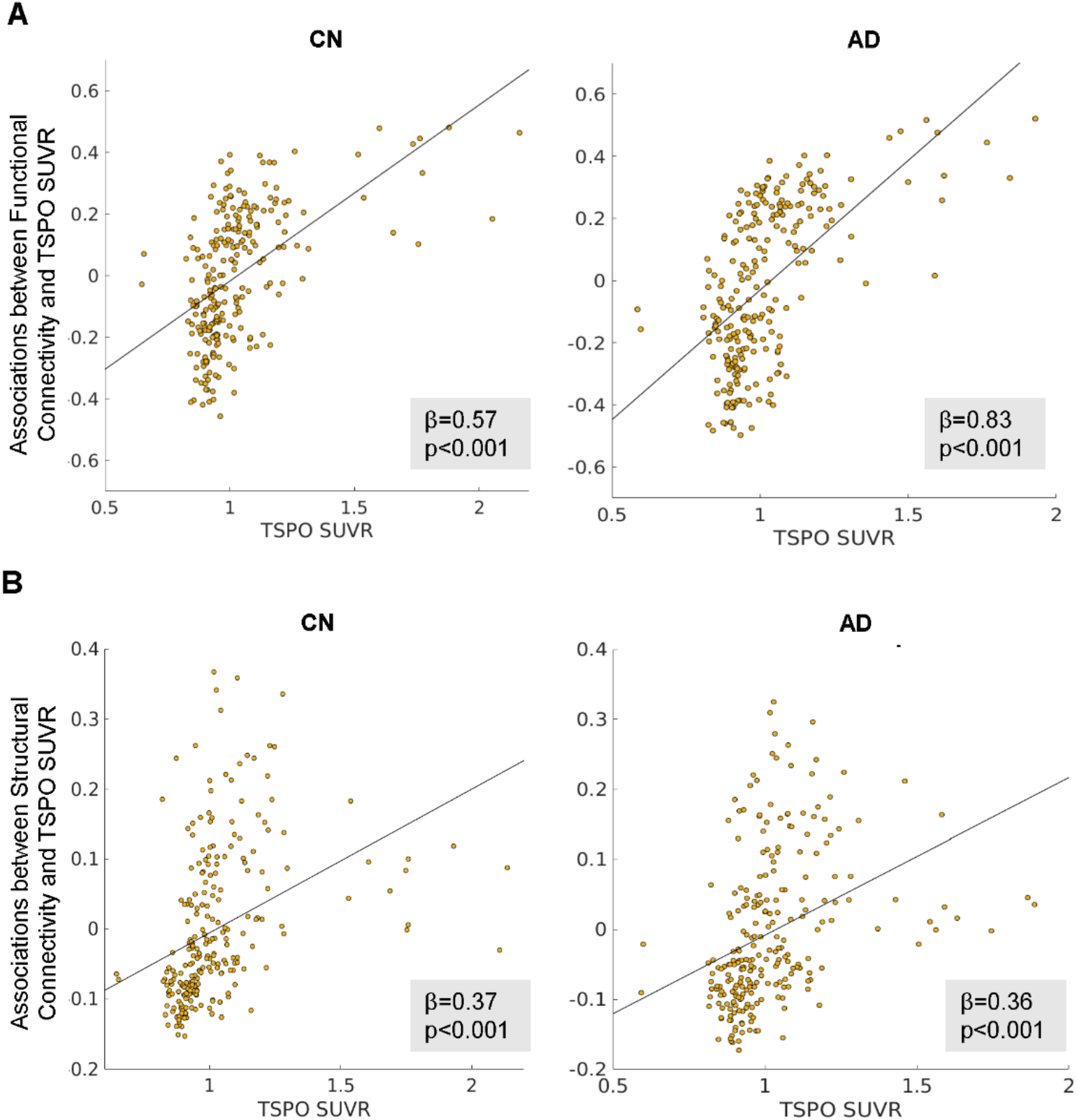
Connectivity is associated with TSPO PET uptake in the target regions depending on TSPO PET uptake in a seed region. **(A)** shows the associators for functional connectivity and **(B)** for structural connectivity. Abbreviations: AD, Alzheimer’s Disease; CN, cognitively normal controls; TSPO SUVR, mitochondrial translocator protein standardized uptake value.

We analyzed how functional connectivity in seed regions with high TSPO PET uptake and in seed regions with low TSPO uptake was associated with the TSPO PET uptake in all target regions. The hot spot seed regions analysis revealed strong positive associations in cognitively normal controls (β=0.79, p<0.001) and AD patients (β=0.74, p<0.001), whereas in the cold spot seed regions analyses a negative association was shown for both groups (cognitively normal controls: β=0.22, p<0.03; AD patients: β=0.25, p=0.002) (**Figure 7A)**. The same analysis based on structural connectivity did not reveal any significant positive or negative associations (**Figure 7B)**.

**Figure 7:**
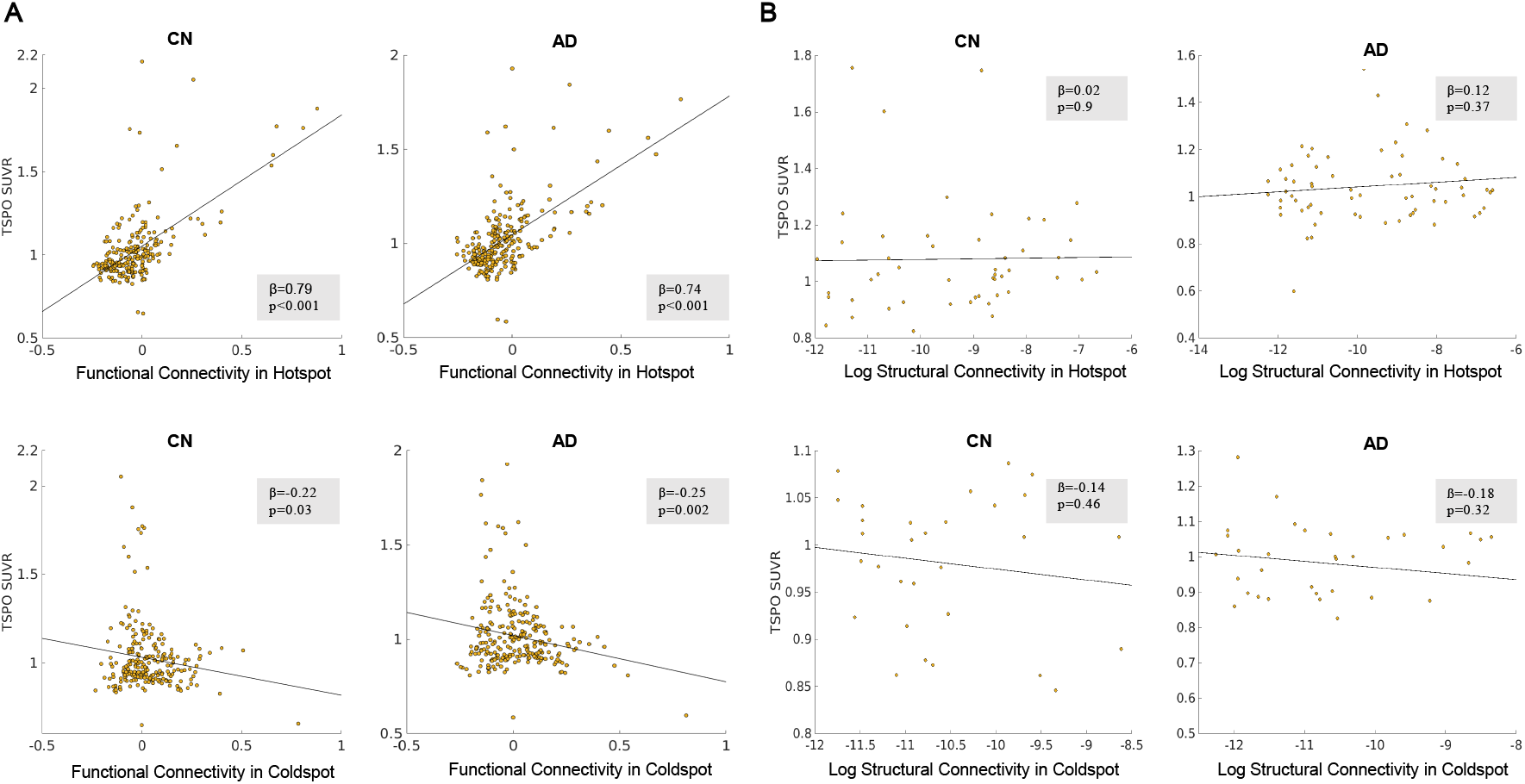
Associations between TSPO PET tracer uptake and (A) functional connectivity in TSPO PET hot spot in temporo-occipital (Brainnetome region 190) and TSPO PET cold spot in the caudate nucleus/basal ganglia (Brainnetome regions 227/228). (B) structural connectivity in TSPO PET hot spot in temporo-occipital (Brainnetome region 190) and TSPO PET cold spot in the caudate nucleus/basal ganglia (Brainnetome region 227/228). Abbreviations: AD, Alzheimer’s Disease; CN, cognitively normal controls; TSPO SUVR, mitochondrial translocator protein standardized uptake value; Log, logarithm.

## Discussion

Microglial activation is increasingly considered the third pathological hallmark of AD, in addition to Aβ and tau (Kinney et al. 2018). The main findings of this study are that (i) in early AD vs cognitively normal controls microglial activation is increased in the temporal lobe (more precisely, the bilateral parahippocampal gyrus), (ii) functional and structural connectivity are reduced globally in AD accompanied by several brain regions showing hyperconnectivity, (iii) microglial activation is associated with dementia severity and cognitive impairment in a disease stage-dependent manner, (iv) functional (and to a lesser degree structural) connectivity is associated with activated microglia and (v) microglial activation spreads along pathways of highly connected brain regions (functional > structural) similar to tau.

Our first finding showed increased parahippocampal microglial activation in AD. This is in line with several previous studies reporting early neurodegenerative and inflammatory changes in the medial temporal lobe (Hamelin et al. 2016; Su et al. 2021; Khan et al. 2014), overlapping with the earliest sites of tau pathology accumulation (Braak et al. 2006). We also confirm the association between dementia severity and cognitive impairment with microglial activation, reported in previous research (Su et al. 2021). The observed quadratic association revealed in the present study indicates a disease stage-dependence of neuroinflammatory response across the AD continuum, earlier suggested for sTREM2, another marker of microglial activation with an inverse u-shaped association between AD severity and CSF sTREM2 levels (Suárez-Calvet et al. 2019; Suárez-Calvet et al. 2016). For TSPO PET this stage dependent association was also observed in a mouse model overexpressing the Aβ precursor protein (Blume et al. 2018). Our study revealed a trend for associations with sTREM2, failing to meet statistical significance due to the relatively small sample size.

Previous research indicates that microglial activation is related to tau propagation and colocalizes with tau pathology in a Braak stage-like fashion; furthermore, activated microglia has been found to precede tau pathology (Hopp et al. 2018; Serrano-Pozo et al. 2011; Pascoal et al. 2021). Although the present study did not explore directly the spatial colocalization of microglial and tau tracer uptake on PET, the Braak stage-like pattern of neuroinflammatory response in early AD still suggest a close relation between both pathologies. In support of this hypothesis a recent multimodal study suggested a strong spatial overlap between tau and microglial PET as well as gray matter atrophy (Su et al. 2021).

The main goal of this study was to assess if functional and structural connectivity are associated with activated microglia. A growing body of evidence supports a model of prion-like tau spreading with progressing neurodegeneration, presumably promoted by neural activity (Franzmeier et al. 2019; Franzmeier et al. 2020; Pereira et al. 2019). In this study, we demonstrated that not only tau, but also microglial activation follows a distribution along functional, and to a lesser degree also structural, connectivity pathways, in line with spatial pathological colocalization. We report that brain areas with higher microglial activation show increased functional connectivity within the same region and with other regions with activated microglia.

We also observed that regions with low microglial activation showed decreased functional connectivity, supporting the hypothesis that not only tau but also activated microglial spread along functionally connected brain areas. Since we observed weaker associations for structural connectivity, one could argue that regions of neuroinflammation are connected by neural activity rather than simply by structural connections.

The described distribution patterns of microglial activation may be of great value for the development of possible anti-inflammatory treatments. They also highlight the importance of an improved mechanistic understanding of spatial spreading of different pathologies across the brain, not limited to Aβ and tau. Strategies to reduce the neural activity-dependent propagation of AD pathology may be a promising avenue for future disease-modifying interventions. It remains to be shown that tau is the leading factor in this process of connectivity-dependent pathology spreading across the brain, followed by a reactive activation of microglia in the same regions. Microglia phagocytose tau and therefore play an important role in spreading of tau pathology throughout the brain (Vogels et al. 2019). There is also evidence that an interaction between microglial activation and Aβ triggers the dissemination of tau across the Braak stages (Pascoal et al. 2021). Further studies including tau and microglia PET should address the question of possible differences in the sequence of tau and inflammation distribution.

There are potential limitations of this study. A relatively small sample size restricts the statistical power of some of the analyses, but previous studies relied on cohorts of comparable size (Völk et al. 2020) and the nature of the presented data is of explorative nature and should be confirmed in independent datasets. No pathological verification was available to confirm the clinical diagnoses; however, a careful biomarker-based stratification and in-depth phenotyping minimizes the likelihood of diagnostic misclassification. Limited blood-brain-barrier passing of [^18^F]GE-180 needs to be acknowledged, but our previous data indicated strong agreement with immunohistochemistry in AD models (<u>Parhizkar et al. 2019</u>) and patterns matching known topology of microglial activation in neurodegenerative diseases (<u>Xiang et al. 2021</u>; Palleis et al. 2021). In fact, the low background signal of [^18^F]GE-180 appears to facilitate sensitive detection of alterations as a specific surrogate of microglial activation (<u>Sridharan et al. 2019</u>; <u>Biechele et al. 2021</u>; Ewers et al. 2020). AD patients were predominantly in an early disease stage, and group differences of microglial activation may have been diluted in the cross-sectional analysis if several activity peaks exist as AD progresses (Fan et al. 2017).

To conclude, the results of this study suggest that microglia-related neuroinflammation in AD spreads along highly connected brain regions. It remains to be explored if this pattern reflects a consequence of trans-neuronal tau spreading or if neuroinflammation itself promotes tau distribution. We confirm that clinical AD is affected by neuroinflammation, and our findings raise the important question if anti-inflammatory treatments could modify trans-neural transmission, emphasizing the need for further comprehensive research integrating mechanisms on the molecular and macro scale brain network levels.

## Data Availability

All data produced in the present study are available upon reasonable request to the authors.

## Acknowledgement

The authors thank Christian Haass, PhD for his support.

This study was supported by the German Center for Neurodegenerative Disorders (Deutsches Zentrum für Neurodegenerative Erkrankungen, DZNE), the Hirnliga e.V. (Manfred-Strohscheer Stiftung) and the Deutsche Forschungsgemeinschaft (DFG, 1007 German Research Foundation) under Germany’s Excellence Strategy within the framework of 1008 the Munich Cluster for Systems Neurology (EXC 2145 SyNergy – ID 390857198).

