## Supplementary Information for "MRI connectivity-based spread of microglial activation in early Alzheimer’s disease"

**Figures**

**Supplementary Figure 1:** Group differences in functional connectivity in the default mode network between early Alzheimer’s disease and cognitively normal controls. Only nodes and edges revealing significant differences after FDR correction are shown.

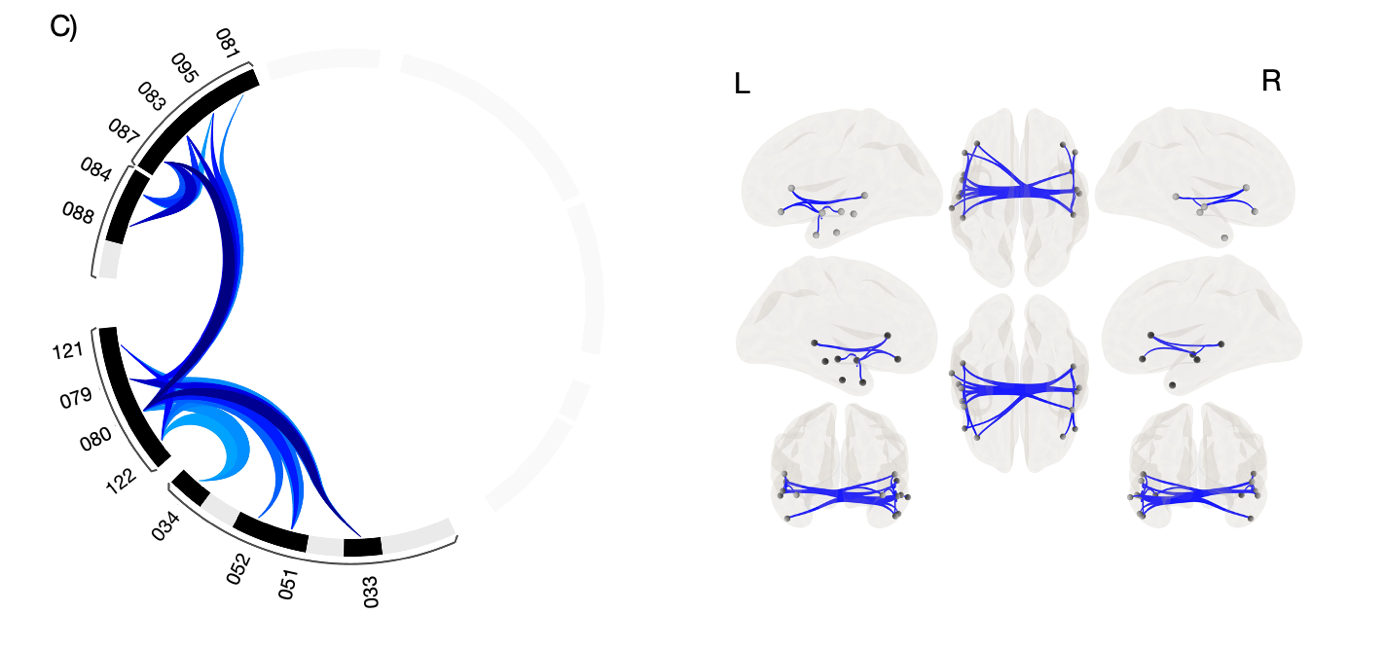

**Tables**

**Supplementary Table 1:** Numbers of Brainnetome regions (https://atlas.brainnetome.org/) with corresponding regions of interest and associated yeo network.

| Label | subregion_name | region | Yeo_7network | Yeo_17network |
| --- | --- | --- | --- | --- |
| 1 | A8m | SFG_L_7_1 | 6 | 17 |
| 2 | A8m | SFG_R_7_1 | 4 | 8 |
| 3 | A8dl | SFG_L_7_2 | 7 | 16 |
| 4 | A8dl | SFG_R_7_2 | 6 | 13 |
| 5 | A9l | SFG_L_7_3 | 7 | 17 |
| 6 | A9l | SFG_R_7_3 | 7 | 17 |
| 7 | A6dl | SFG_L_7_4 | 3 | 6 |
| 8 | A6dl | SFG_R_7_4 | 3 | 6 |
| 9 | A6m | SFG_L_7_5 | 2 | 3 |
| 10 | A6m | SFG_R_7_5 | 2 | 3 |
| 11 | A9m | SFG_L_7_6 | 7 | 13 |
| 12 | A9m | SFG_R_7_6 | 6 | 13 |
| 13 | A10m | SFG_L_7_7 | 7 | 16 |
| 14 | A10m | SFG_R_7_7 | 7 | 16 |
| 15 | A9/46d | MFG_L_7_1 | 4 | 8 |
| 16 | A9/46d | MFG_R_7_1 | 6 | 11 |
| 17 | IFJ | MFG_L_7_2 | 6 | 12 |
| 18 | IFJ | MFG_R_7_2 | 6 | 12 |
| 19 | A46 | MFG_L_7_3 | 6 | 13 |
| 20 | A46 | MFG_R_7_3 | 6 | 13 |
| 21 | A9/46v | MFG_L_7_4 | 6 | 12 |
| 22 | A9/46v | MFG_R_7_4 | 6 | 8 |
| 23 | A8vl | MFG_L_7_5 | 7 | 16 |
| 24 | A8vl | MFG_R_7_5 | 6 | 13 |
| 25 | A6vl | MFG_L_7_6 | 3 | 6 |
| 26 | A6vl | MFG_R_7_6 | 3 | 12 |
| 27 | A10l | MFG_L_7_7 | 5 | 10 |
| 28 | A10l | MFG_R_7_7 | 6 | 13 |
| 29 | A44d | IFG_L_6_1 | 6 | 12 |
| 30 | A44d | IFG_R_6_1 | 3 | 12 |
| 31 | IFS | IFG_L_6_2 | 6 | 12 |
| 32 | IFS | IFG_R_6_2 | 6 | 12 |
| 33 | A45c | IFG_L_6_3 | 7 | 17 |
| 34 | A45c | IFG_R_6_3 | 7 | 17 |
| 35 | A45r | IFG_L_6_4 | 7 | 17 |
| 36 | A45r | IFG_R_6_4 | 6 | 8 |
| 37 | A44op | IFG_L_6_5 | 4 | 7 |
| 38 | A44op | IFG_R_6_5 | 4 | 8 |
| 39 | A44v | IFG_L_6_6 | 4 | 8 |
| 40 | A44v | IFG_R_6_6 | 4 | 8 |
| 41 | A14m | OrG_L_6_1 | 7 | 16 |
| 42 | A14m | OrG_R_6_1 | 7 | 16 |
| 43 | A12/47o | OrG_L_6_2 | 7 | 17 |
| 44 | A12/47o | OrG_R_6_2 | 7 | 17 |
| 45 | A11l | OrG_L_6_3 | 5 | 10 |
| 46 | A11l | OrG_R_6_3 | 6 | 12 |
| 47 | A11m | OrG_L_6_4 | 5 | 10 |
| 48 | A11m | OrG_R_6_4 | 5 | 10 |
| 49 | A13 | OrG_L_6_5 | 5 | 10 |
| 50 | A13 | OrG_R_6_5 | 5 | 10 |
| 51 | A12/47l | OrG_L_6_6 | 7 | 17 |
| 52 | A12/47l | OrG_R_6_6 | 7 | 17 |
| 53 | A4hf | PrG_L_6_1 | 2 | 4 |
| 54 | A4hf | PrG_R_6_1 | 2 | 4 |
| 55 | A6cdl | PrG_L_6_2 | 3 | 6 |
| 56 | A6cdl | PrG_R_6_2 | 3 | 6 |
| 57 | A4ul | PrG_L_6_3 | 2 | 3 |
| 58 | A4ul | PrG_R_6_3 | 2 | 3 |
| 59 | A4t | PrG_L_6_4 | 2 | 3 |
| 60 | A4t | PrG_R_6_4 | 2 | 3 |
| 61 | A4tl | PrG_L_6_5 | 4 | 7 |
| 62 | A4tl | PrG_R_6_5 | 4 | 7 |
| 63 | A6cvl | PrG_L_6_6 | 3 | 12 |
| 64 | A6cvl | PrG_R_6_6 | 3 | 12 |
| 65 | A1/2/3ll | PCL_L_2_1 | 4 | 7 |
| 66 | A1/2/3ll | PCL_R_2_1 | 2 | 3 |
| 67 | A4ll | PCL_L_2_2 | 2 | 3 |
| 68 | A4ll | PCL_R_2_2 | 2 | 3 |
| 69 | A38m | STG_L_6_1 | 5 | 9 |
| 70 | A38m | STG_R_6_1 | 5 | 9 |
| 71 | A41/42 | STG_L_6_2 | 2 | 4 |
| 72 | A41/42 | STG_R_6_2 | 2 | 4 |
| 73 | TE1.0/TE1.2 | STG_L_6_3 | 2 | 4 |
| 74 | TE1.0/TE1.2 | STG_R_6_3 | 2 | 4 |
| 75 | A22c | STG_L_6_4 | 2 | 14 |
| 76 | A22c | STG_R_6_4 | 2 | 4 |
| 77 | A38l | STG_L_6_5 | 5 | 9 |
| 78 | A38l | STG_R_6_5 | 5 | 9 |
| 79 | A22r | STG_L_6_6 | 7 | 14 |
| 80 | A22r | STG_R_6_6 | 7 | 17 |
| 81 | A21c | MTG_L_4_1 | 7 | 17 |
| 82 | A21c | MTG_R_4_1 | 6 | 13 |
| 83 | A21r | MTG_L_4_2 | 7 | 17 |
| 84 | A21r | MTG_R_4_2 | 7 | 17 |
| 85 | A37dl | MTG_L_4_3 | 3 | 6 |
| 86 | A37dl | MTG_R_4_3 | 3 | 6 |
| 87 | aSTS | MTG_L_4_4 | 7 | 17 |
| 88 | aSTS | MTG_R_4_4 | 7 | 14 |
| 89 | A20iv | ITG_L_7_1 | 5 | 9 |
| 90 | A20iv | ITG_R_7_1 | 5 | 9 |
| 91 | A37elv | ITG_L_7_2 | 3 | 5 |
| 92 | A37elv | ITG_R_7_2 | 3 | 5 |
| 93 | A20r | ITG_L_7_3 | 5 | 9 |
| 94 | A20r | ITG_R_7_3 | 5 | 9 |
| 95 | A20il | ITG_L_7_4 | 7 | 17 |
| 96 | A20il | ITG_R_7_4 | 5 | 17 |
| 97 | A37vl | ITG_L_7_5 | 3 | 12 |
| 98 | A37vl | ITG_R_7_5 | 3 | 5 |
| 99 | A20cl | ITG_L_7_6 | 6 | 13 |
| 100 | A20cl | ITG_R_7_6 | 6 | 13 |
| 101 | A20cv | ITG_L_7_7 | 5 | 9 |
| 102 | A20cv | ITG_R_7_7 | 5 | 9 |
| 103 | A20rv | FuG_L_3_1 | 5 | 9 |
| 104 | A20rv | FuG_R_3_1 | 5 | 9 |
| 105 | A37mv | FuG_L_3_2 | 1 | 1 |
| 106 | A37mv | FuG_R_3_2 | 1 | 1 |
| 107 | A37lv | FuG_L_3_3 | 3 | 5 |
| 108 | A37lv | FuG_R_3_3 | 1 | 5 |
| 109 | A35/36r | PhG_L_6_1 | 5 | 9 |
| 110 | A35/36r | PhG_R_6_1 | 5 | 9 |
| 111 | A35/36c | PhG_L_6_2 | 5 | 15 |
| 112 | A35/36c | PhG_R_6_2 | 1 | 15 |
| 113 | TL | PhG_L_6_3 | 1 | 15 |
| 114 | TL | PhG_R_6_3 | 1 | 15 |
| 115 | A28/34 | PhG_L_6_4 | 5 | 9 |
| 116 | A28/34 | PhG_R_6_4 | 5 | 9 |
| 117 | TI | PhG_L_6_5 | 5 | 9 |
| 118 | TI | PhG_R_6_5 | 5 | 9 |
| 119 | TH | PhG_L_6_6 | 1 | 2 |
| 120 | TH | PhG_R_6_6 | 1 | 2 |
| 121 | rpSTS | pSTS_L_2_1 | 7 | 14 |
| 122 | rpSTS | pSTS_R_2_1 | 7 | 14 |
| 123 | cpSTS | pSTS_L_2_2 | 4 | 14 |
| 124 | cpSTS | pSTS_R_2_2 | 4 | 14 |
| 125 | A7r | SPL_L_5_1 | 3 | 6 |
| 126 | A7r | SPL_R_5_1 | 3 | 6 |
| 127 | A7c | SPL_L_5_2 | 3 | 5 |
| 128 | A7c | SPL_R_5_2 | 3 | 5 |
| 129 | A5l | SPL_L_5_3 | 3 | 6 |
| 130 | A5l | SPL_R_5_3 | 3 | 6 |
| 131 | A7pc | SPL_L_5_4 | 2 | 3 |
| 132 | A7pc | SPL_R_5_4 | 2 | 3 |
| 133 | A7ip | SPL_L_5_5 | 3 | 5 |
| 134 | A7ip | SPL_R_5_5 | 3 | 5 |
| 135 | A39c | IPL_L_6_1 | 1 | 5 |
| 136 | A39c | IPL_R_6_1 | 1 | 5 |
| 137 | A39rd | IPL_L_6_2 | 6 | 12 |
| 138 | A39rd | IPL_R_6_2 | 6 | 12 |
| 139 | A40rd | IPL_L_6_3 | 3 | 6 |
| 140 | A40rd | IPL_R_6_3 | 3 | 6 |
| 141 | A40c | IPL_L_6_4 | 7 | 17 |
| 142 | A40c | IPL_R_6_4 | 6 | 13 |
| 143 | A39rv | IPL_L_6_5 | 3 | 14 |
| 144 | A39rv | IPL_R_6_5 | 7 | 16 |
| 145 | A40rv | IPL_L_6_6 | 2 | 4 |
| 146 | A40rv | IPL_R_6_6 | 2 | 4 |
| 147 | A7m | PCun_L_4_1 | 6 | 11 |
| 148 | A7m | PCun_R_4_1 | 6 | 11 |
| 149 | A5m | PCun_L_4_2 | 2 | 3 |
| 150 | A5m | PCun_R_4_2 | 3 | 6 |
| 151 | dmPOS | PCun_L_4_3 | 1 | 2 |
| 152 | dmPOS | PCun_R_4_3 | 1 | 2 |
| 153 | A31 | PCun_L_4_4 | 7 | 16 |
| 154 | A31 | PCun_R_4_4 | 7 | 16 |
| 155 | A1/2/3ulhf | PoG_L_4_1 | 2 | 4 |
| 156 | A1/2/3ulhf | PoG_R_4_1 | 2 | 4 |
| 157 | A1/2/3tonIa | PoG_L_4_2 | 2 | 4 |
| 158 | A1/2/3tonIa | PoG_R_4_2 | 2 | 4 |
| 159 | A2 | PoG_L_4_3 | 3 | 6 |
| 160 | A2 | PoG_R_4_3 | 2 | 3 |
| 161 | A1/2/3tru | PoG_L_4_4 | 2 | 3 |
| 162 | A1/2/3tru | PoG_R_4_4 | 2 | 3 |
| 163 | G | INS_L_6_1 | 2 | 4 |
| 164 | G | INS_R_6_1 | 2 | 4 |
| 165 | vIa | INS_L_6_2 | 0 | 0 |
| 166 | vIa | INS_R_6_2 | 6 | 8 |
| 167 | dIa | INS_L_6_3 | 4 | 8 |
| 168 | dIa | INS_R_6_3 | 4 | 8 |
| 169 | vId/vIg | INS_L_6_4 | 4 | 7 |
| 170 | vId/vIg | INS_R_6_4 | 4 | 7 |
| 171 | dIg | INS_L_6_5 | 2 | 4 |
| 172 | dIg | INS_R_6_5 | 2 | 4 |
| 173 | dId | INS_L_6_6 | 4 | 7 |
| 174 | dId | INS_R_6_6 | 4 | 7 |
| 175 | A23d | CG_L_7_1 | 7 | 16 |
| 176 | A23d | CG_R_7_1 | 7 | 16 |
| 177 | A24rv | CG_L_7_2 | 0 | 0 |
| 178 | A24rv | CG_R_7_2 | 0 | 0 |
| 179 | A32p | CG_L_7_3 | 7 | 16 |
| 180 | A32p | CG_R_7_3 | 4 | 8 |
| 181 | A23v | CG_L_7_4 | 7 | 15 |
| 182 | A23v | CG_R_7_4 | 1 | 15 |
| 183 | A24cd | CG_L_7_5 | 4 | 7 |
| 184 | A24cd | CG_R_7_5 | 4 | 7 |
| 185 | A23c | CG_L_7_6 | 4 | 8 |
| 186 | A23c | CG_R_7_6 | 4 | 7 |
| 187 | A32sg | CG_L_7_7 | 7 | 16 |
| 188 | A32sg | CG_R_7_7 | 7 | 16 |
| 189 | cLinG | MVOcC_L_5_1 | 1 | 1 |
| 190 | cLinG | MVOcC_R_5_1 | 1 | 1 |
| 191 | rCunG | MVOcC_L_5_2 | 1 | 2 |
| 192 | rCunG | MVOcC_R_5_2 | 1 | 2 |
| 193 | cCunG | MVOcC_L_5_3 | 1 | 1 |
| 194 | cCunG | MVOcC_R_5_3 | 1 | 1 |
| 195 | rLinG | MVOcC_L_5_4 | 1 | 2 |
| 196 | rLinG | MVOcC_R_5_4 | 1 | 1 |
| 197 | vmPOS | MVOcC_L_5_5 | 1 | 2 |
| 198 | vmPOS | MVOcC_R_5_5 | 1 | 2 |
| 199 | mOccG | LOcC_L_4_1 | 1 | 1 |
| 200 | mOccG | LOcC_R_4_1 | 1 | 1 |
| 201 | V5/MT+ | LOcC_L_4_2 | 3 | 5 |
| 202 | V5/MT+ | LOcC_R_4_2 | 1 | 5 |
| 203 | OPC | LOcC_L_4_3 | 1 | 1 |
| 204 | OPC | LOcC_R_4_3 | 1 | 1 |
| 205 | iOccG | LOcC_L_4_4 | 1 | 1 |
| 206 | iOccG | LOcC_R_4_4 | 1 | 1 |
| 207 | msOccG | LOcC_L_2_1 | 1 | 2 |
| 208 | msOccG | LOcC_R_2_1 | 1 | 2 |
| 209 | lsOccG | LOcC_L_2_2 | 1 | 1 |
| 210 | lsOccG | LOcC_R_2_2 | 1 | 5 |
| 211 | mAmyg | Amyg_L_2_1 | 0 | 0 |
| 212 | mAmyg | Amyg_R_2_1 | 0 | 0 |
| 213 | lAmyg | Amyg_L_2_2 | 0 | 0 |
| 214 | lAmyg | Amyg_R_2_2 | 0 | 0 |
| 215 | rHipp | Hipp_L_2_1 | 0 | 0 |
| 216 | rHipp | Hipp_R_2_1 | 0 | 0 |
| 217 | cHipp | Hipp_L_2_2 | 0 | 0 |
| 218 | cHipp | Hipp_R_2_2 | 0 | 0 |
| 219 | vCa | BG_L_6_1 | 0 | 0 |
| 220 | vCa | BG_R_6_1 | 0 | 0 |
| 221 | GP | BG_L_6_2 | 0 | 0 |
| 222 | GP | BG_R_6_2 | 0 | 0 |
| 223 | NAC | BG_L_6_3 | 0 | 0 |
| 224 | NAC | BG_R_6_3 | 0 | 0 |
| 225 | vmPu | BG_L_6_4 | 0 | 0 |
| 226 | vmPu | BG_R_6_4 | 0 | 0 |
| 227 | dCa | BG_L_6_5 | 0 | 0 |
| 228 | dCa | BG_R_6_5 | 0 | 0 |
| 229 | dlPu | BG_L_6_6 | 0 | 0 |
| 230 | dlPu | BG_R_6_6 | 0 | 0 |
| 231 | mPFtha | Tha_L_8_1 | 0 | 0 |
| 232 | mPFtha | Tha_R_8_1 | 0 | 0 |
| 233 | mPMtha | Tha_L_8_2 | 0 | 0 |
| 234 | mPMtha | Tha_R_8_2 | 0 | 0 |
| 235 | Stha | Tha_L_8_3 | 0 | 0 |
| 236 | Stha | Tha_R_8_3 | 0 | 0 |
| 237 | rTtha | Tha_L_8_4 | 0 | 0 |
| 238 | rTtha | Tha_R_8_4 | 0 | 0 |
| 239 | PPtha | Tha_L_8_5 | 0 | 0 |
| 240 | PPtha | Tha_R_8_5 | 0 | 0 |
| 241 | Otha | Tha_L_8_6 | 0 | 0 |
| 242 | Otha | Tha_R_8_6 | 0 | 0 |
| 243 | cTtha | Tha_L_8_7 | 0 | 0 |
| 244 | cTtha | Tha_R_8_7 | 0 | 0 |
| 245 | lPFtha | Tha_L_8_8 | 0 | 0 |
| 246 | lPFtha | Tha_R_8_8 | 0 | 0 |
